# Emergence and predominance of novel influenza A (H3N2) subclade (3C.2a1b.2a.2a.3a.1) in Kenya and globally in 2023

**DOI:** 10.1101/2023.12.15.23299920

**Authors:** John Mwita Morobe, Edidah Moraa, Arnold W. Lambisia, Martin Mutunga, Brenda Kiage, Esther Nyadzua Katama, Timothy Makori, Joyce Nyiro, Leonard Ndwiga, James Nyagwange, Charles Sande, Lynette Isabella Ochola-Oyier, Edward C. Holmes, D. James Nokes, George Githinji, Charles N. Agoti

**Affiliations:** Kenya Medical Research Institute Wellcome Trust Research Programme, Kilifi, Kenya; The University of Sydney; University of Warwick, Coventry, UK; Pwani University, Kilifi

## Abstract

We report emergence and predominance of the influenza A (H3N2) subclade 3C.2a1b.2a.2a.3a.1 in Kenya similar to the global clade in 2023. The Kenyan 3C.2a1b.2a.2a.3a.1 viruses have >15 amino acid differences in the HA and NA proteins relative to 2023/24 WHO recommended Northern/Southern Hemisphere influenza vaccine strains.

## Main

Antigenic drift in the hemagglutinin (HA) and neuraminidase (NA) proteins of influenza A/B viruses is a key contributor to their recurrent global epidemics^1^. Influenza genomic surveillance helps identify virus strains likely to predominate in future epidemics, enabling appropriate vaccine strain recommendations^2^. Critically, however, the limited influenza genomic surveillance in sub-Saharan Africa obscures global influenza epidemiology.

The KEMRI-Wellcome Trust Research Programme, coastal Kenya, runs two respiratory pathogen surveillance platforms: (a) paediatric inpatient surveillance of severe and very severe pneumonia in <5-year-olds at Kilifi County Hospital^3^ and (b) all age groups outpatient acute respiratory infection (ARI) surveillance at five Kilifi County health facilities (HF)^4^. The former surveillance collects respiratory samples from all eligible consented patients, while the latter collects ∼15 samples per HF per week. Influenza infection is diagnosed using RT-PCR^5^.

Between 1st January and 29th September 2023, 3,262 respiratory samples were collected from the two platforms^6^. Only 1,192 (37%) of collected samples were screened due to reagent limitations. Of these, 6% (68/1192) were positive for influenza A (n=58, 85%), B (n=6, 9%) or C (n=4, 6%) viruses; The weekly influenza A/B/C detections are shown in **Figure 1A**.

**Figure 1.**
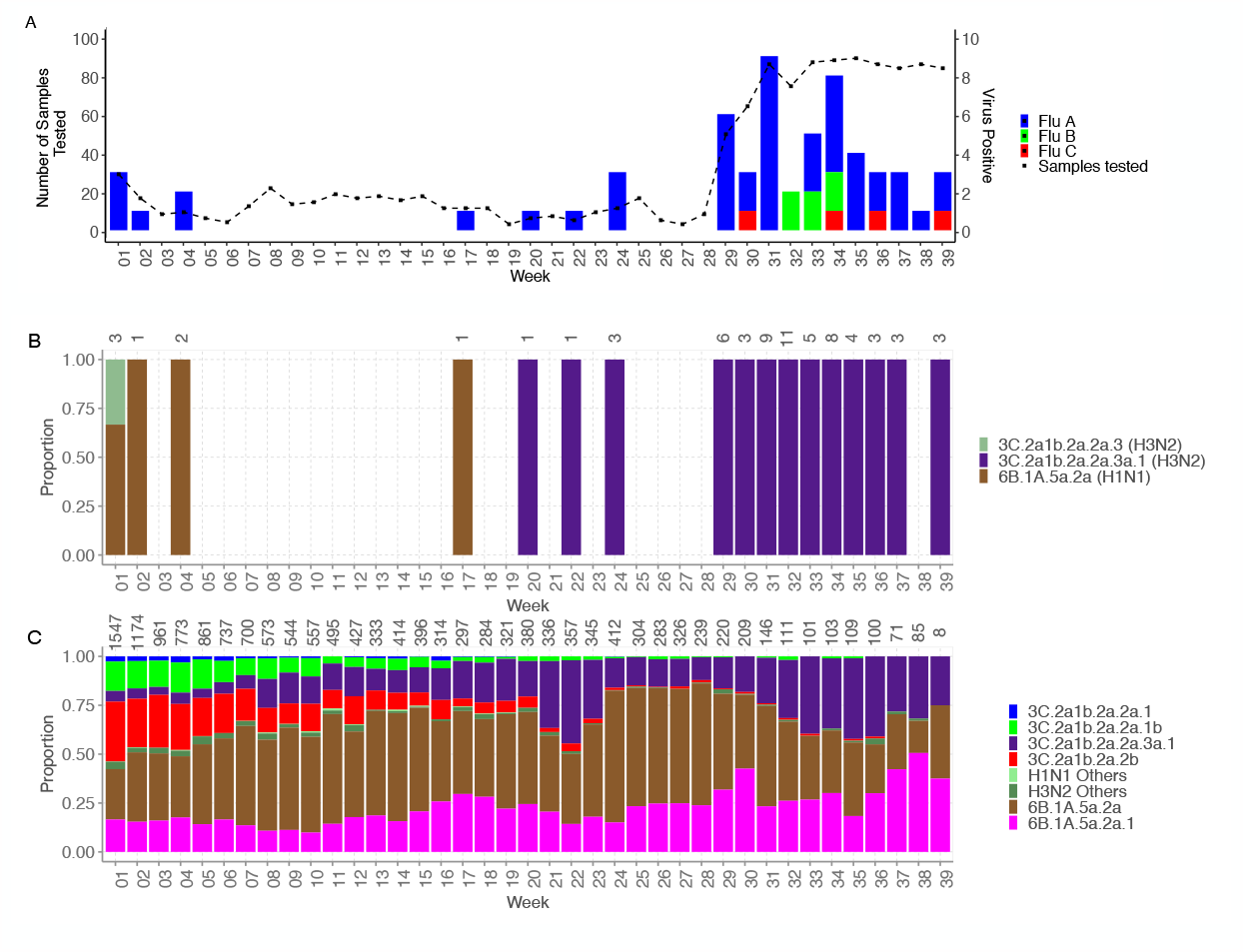
Temporal distribution of influenza in Kenya and globally. A) Weekly detections of influenza virus type A, B and C in samples collected in Kilifi between 1st January and 29th September 2023. The observed low testing numbers (10-20 samples per week) up to week 28, followed by a rapid increase to 80 per week is attributed to a batch of samples that were not tested during weeks 0 to 28 due to reagents constraints. B) Distribution of clades/subclades within the influenza subtype A(H3N2) pdm09 and A(H1N1) subtype in Kilifi. The numbers on top of the bars indicate the number of sequences available for each analyzed week. C) Distribution of clades/subclades within the influenza subtype A(H3N2) pdm09 and A(H1N1) subtype worldwide in 2023 (Jan-Sep) based on data deposited on GISAID (accessed on 7th November 2023).

Influenza A positives were targeted for a multi-segment PCR^7^ and sequencing on Illumina MiSeq or ONT GridION. Raw reads were assembled using the reference-based FLU or FLU-MinION module of the Iterative Refinement Meta-Assembler (IRMA)^8^. Recovered sequences were assigned phylogenetic clades /subclades based on HA gene using NextClade^9^. Maximum likelihood (ML) phylogenetic trees were estimated using IQ-TREE v2.2.2.6^10^, and time-scaled phylogenies were estimated using TimeTree^11^.

Full-length viral genomes (>90% coverage) were recovered from 52 of 58 samples and deposited in GISAID EpiFlu™ (accession nos EPI_ISL_18560856 - EPI_ISL_18560859 and EPI_ISL_18560907 - EPI_ISL_18560947). These were classified as A(H1N1) pdm09 (n=6, 12%) and A(H3N2) (n=46, 88%). Three clades/subclades were identified: one within A(H1N1) pdm09 and two within A(H3N2). The (H3N2) 3C.2a1b.2a.2a.3a.1 viruses, first seen in the Kenya on 19th May 2023, displaced previously circulating clades/subclades within the A(H3N2) subtype (**Figure 1B)**. Based on GISAID data, the 3C.2a1b.2a.2a.3a.1 viruses were first detected in United Arab Emirates in January 2022, and have displaced other A(H3N2) clades/sub-clades globally (**Figure 1C**). The Kenya patients infected with 3C.2a1b.2a.2a.3a.1 viruses presented with cough (100%), fever (82%), nasal discharge (80%) and difficulty breathing (13%) **(Table 1)**.

**Table 1.** Signs and symptoms present among patients infected with influenza A (H3N2) 3C.2a1b.2a.2a.3a.1 viruses, Kilifi, Kenya, January–September 2023.

| Signs/symptoms | No. (%) patients (n=45) |
| --- | --- |
| **Fever** |  |
| Yes | 37 (82%) |
| **Cough** |  |
| Yes | 45 (100%) |
| **Nasal discharge** |  |
| Yes | 36 (80%) |
| **Difficulty breathing** |  |
| Yes | 6 (13%) |
| **Sore throat** |  |
| Yes | 3 (7%) |
| **Body malaise** |  |
| Yes | 10 (22%) |
| **Chest indrawing** |  |
| Yes | 0 (0%) |
| **Wheezing** |  |
| Yes | 1 (2%) |

The global phylogeny of the 3C.2a1b.2a.2a.3a.1 subclade (n=3,462) revealed that the Kenyan sequences fall into 4 groups (two clusters, named C1 and C2, and 2 singletons, named S1 and S2) indicating at least 4 separate introductions into the country **(Appendix 1 Figure 1)**. The Kenya HA and NA sequences differed from the global sequences at multiple nucleotide positions: NA (G435A, A687G, A1230G and C1275T) and HA (deletion at 131, G991A, A1035G and G1124A). Based on the HA gene segment, the mean time to the most recent common ancestor for the 3C.2a1b.2a.2a.3a.1 subclade viruses was estimated to be April 2019 [95% Highest Posterior Density (HPD): August 2018 – January 2020], with a nucleotide substitution rate of 1.19×10-3 [95% HPD: 9.53 × 10-4 – 1.40 × 10-3] substitutions/site/year.

There were amino acid changes between the HA of the Kenyan 3C.2a1b.2a.2a.3a.1 viruses and the 2023/24 season WHO recommended vaccine strains: 26 and 27 amino acid changes relative to the HA protein of the Northern Hemisphere (NH) vaccine strains – A/Darwin/6/2021 (H3N2)-like virus and A/Darwin/9/2021(H3N2)-like virus, respectively **(Appendix 1 Figure 2A and B)**. The substitutions included T3A (signal peptide), E50K, D53N, G53N, N96S, I140K, N186D, I192F, I223V, G225D in HA1, and N49S (HA2). These mutations were seen in 100% of the Kenya viruses. There were 17 amino acid changes relative to the HA of the Southern Hemisphere (SH) vaccine strain – A/Massachusetts/18/2022 (H3N2)-like virus, **(Appendix 1 Figure 2C)**. Substitutions included I58V(HA1) observed in 11% of the viruses, V77I (HA2) observed in 8% of the viruses. Comparison of the Kenya viruses to the SH vaccine strain A/Thailand/18/2022 (H3N2)-like virus also revealed 17 amino acid changes **(Appendix 1 Figure 2D)** with the most frequent HA substitution being I58V(HA1) observed in 11% of the viruses, V77I (HA2) observed in 8% of the viruses.

Relative to the NA protein of Northern Hemisphere (NH) vaccine strains A/Darwin/6/2021 (H3N2)-like virus and A/Darwin/9/2021(H3N2)-like virus, showed 22 and 21 amino acid changes, respectively **(Appendix 1 Figure 3A and B**). A comparison with SH vaccine strain A/Thailand/18/2022 (H3N2)-like virus and A/Massachusetts/18/2022 (H3N2)-like virus revealed 21 and 20 amino acid changes respectively in NA protein **(Appendix 1 Figure 3C and D)**.

The factors contributing to the global emergence and predominance of 3C.2a1b.2a.2a.3a.1 viruses over other A(H3N2) strains are currently unclear. We speculate that the H1 mutations observed in this subclade likely enhance fitness over the other subclades^12,13^, or make it less immunologically recognizable to current immunity established by vaccination or natural infection^14^. Our findings emphasize the need for near-real time genomic surveillance globally, including LMICs, to monitor the evolution of circulating IAV for optimal vaccine strain selection.

## Data Availability

All data generated and analysis script for this manuscript are available
from the Virus Epidemiology and Control, Kenya Medical Research Institute (KEMRI)Wellcome
Trust Research Programme data server:https://doi.org/10.7910/DVN/5US8MM, Full-length viral genomes are available in GISAID EpiFlu (accession nos EPI_ISL_18560856 - EPI_ISL_18560859 and EPI_ISL_18560907 - EPI_ISL_18560947)

## Acknowledgments

We thank the members of the Viral Epidemiology and Control Research Group at KEMRI Wellcome Trust Research Programme, field, clinical and laboratory staff, for their valuable contributions to this work. We also thank laboratories that generated and shared genetic sequence data via the GISAID Initiative (https://www.gisaid.org), on which this part of this research is based. Genome sequences generated in this study are available in the GISAID’s EpiFlu database (accession nos EPI_ISL_18560856 - EPI_ISL_18560859 and EPI_ISL_18560907 -EPI_ISL_18560947). The dataset and analysis scripts used are available in the Harvard Dataverse repository (https://doi.org/10.7910/DVN/5US8MM).

## Figure and Table

**Appendix 1 Figure 1.**
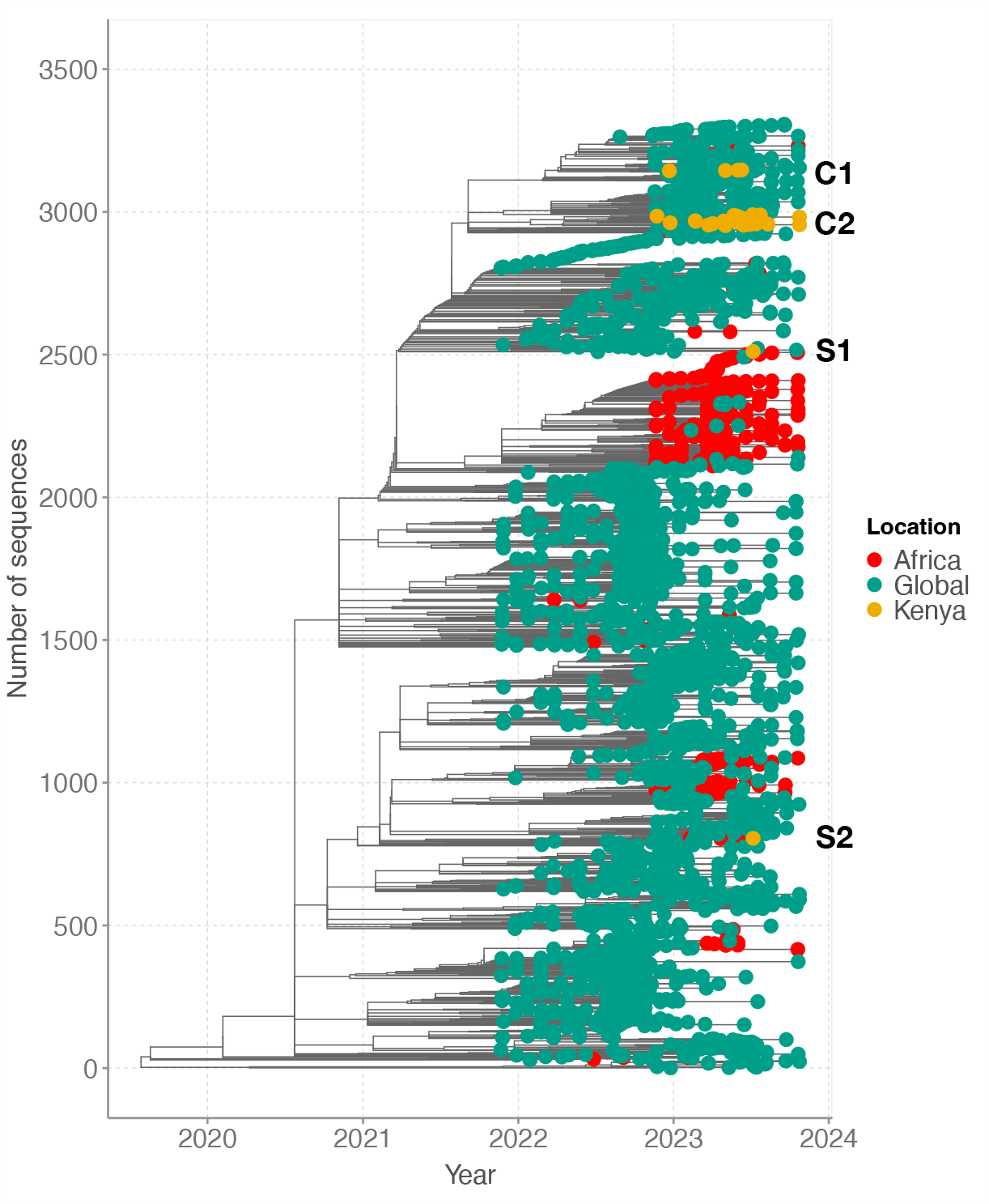
Time-resolved phylogeny of global genomes(n=3,462) combined with Kilifi IAV genomes (n=45). Global genomes are depicted in green, African genomes in red, and Kenyan genomes in orange.

**Appendix 1 Figure 2.**
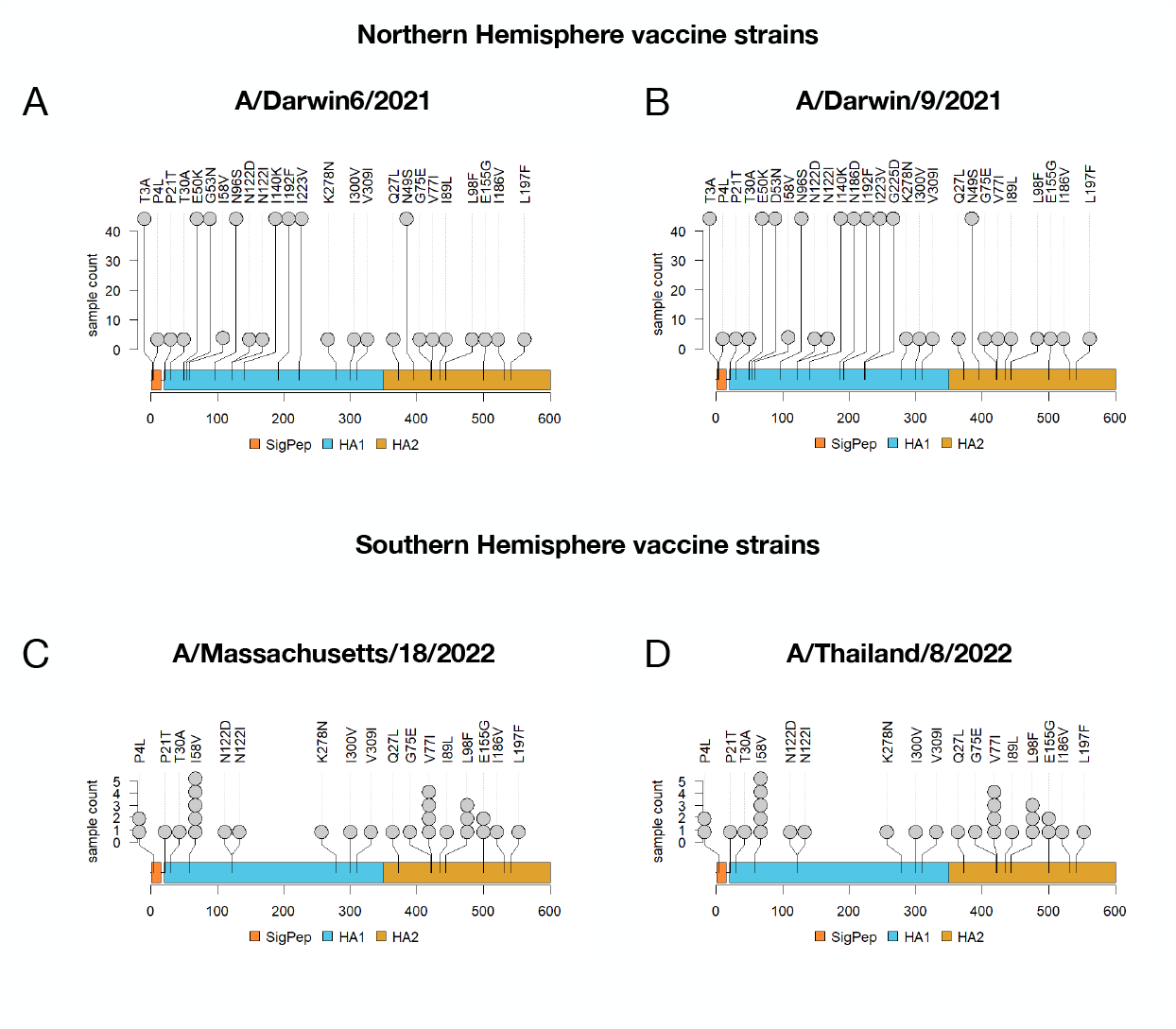
Amino acid sequence differences in the HA protein. The lollipop plot represents the positions and amino acid difference in the HA protein in comparison to the respective Northern Hemisphere (NH) and Southern Hemisphere (SH) WHO recommended vaccine strains.

**Appendix 1 Figure 3.**
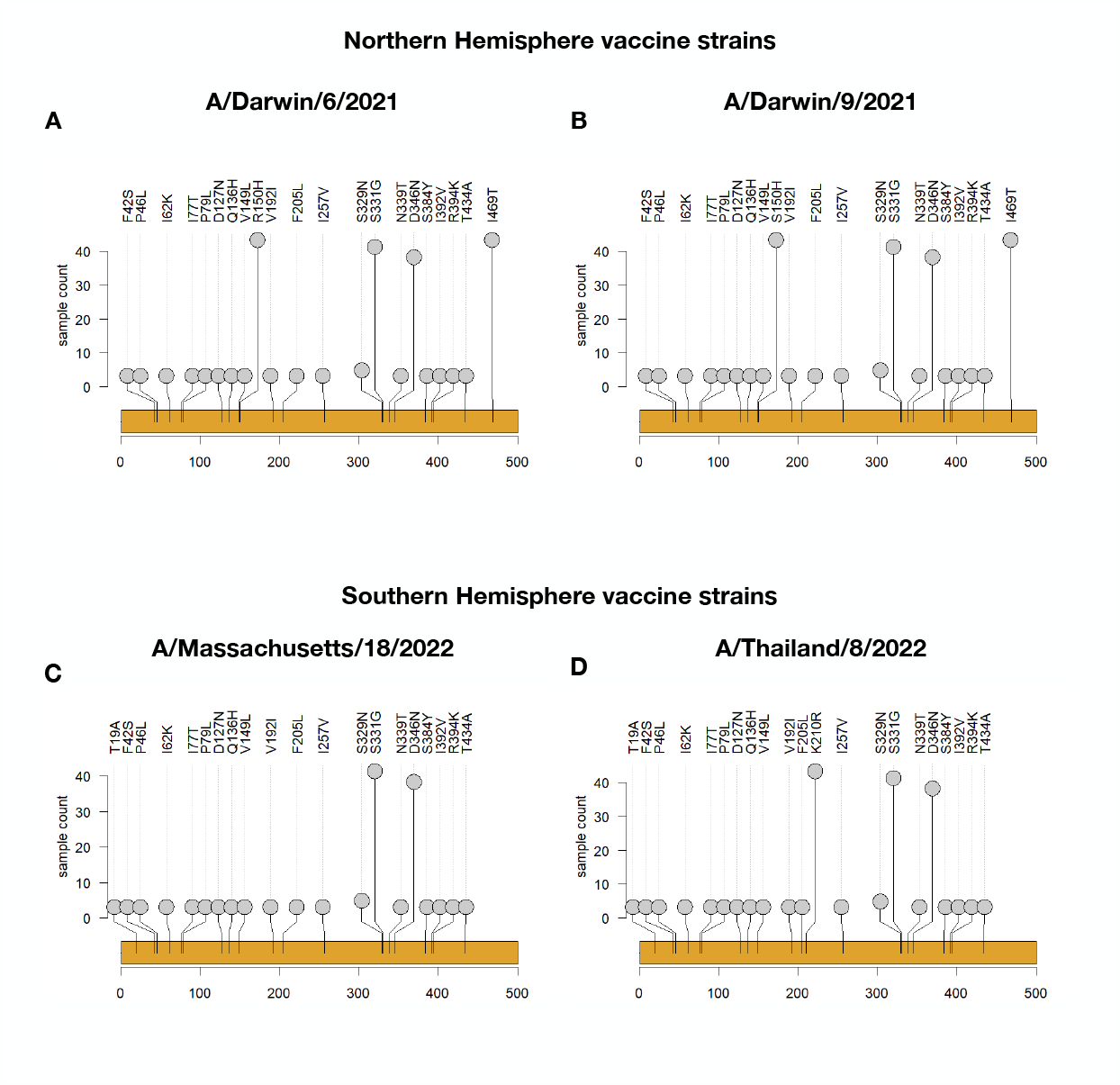
Amino acid sequence differences in the NA protein. The lollipop plot represents the positions and amino acid difference in the NA protein in comparison to the respective Northern Hemisphere (NH) and Southern Hemisphere (SH) WHO recommended vaccine strains.

